# A first study on the impact of containment measure on COVID-19 spread in Morocco

**DOI:** 10.1101/2020.04.26.20080770

**Authors:** Aayah Hammoumi, Redouane Qesmi

**Affiliations:** Department of Biology, Cadi Ayyad University, Semlalia, Marrakech 40000, Morocco; Superior School of Technology, Sidi Mohamed Ben Abdellah University, Fez 30000, Morocco

**Keywords:** Covid-19, Containment, Asymptomatic, Symptomatic, Unreported, Epidemiological model, Basic reproduction number.

## Abstract

**Background:** Since the appearance of the first case of COVID-19 in Morocco, the cumulative number of reported infectious cases continues to increase and, consequently, the government imposed the containment measure within the country. Our aim is to predict the impact of the compulsory containment on COVID-19 spread. Earlier knowledge of the epidemic characteristics of COVID-19 transmission related to Morocco will be of great interest to establish an optimal plan-of-action to control the epidemic.

**Method:** Using a Susceptible-Asymptomatic-Infectious model and the data of reported cumulative confirmed cases in Morocco from March 2nd to April 9, 2020, we determined the basic and control reproduction numbers and we estimated the model parameter values. Furthermore, simulations of different scenarios of containment are performed.

**Results:** Epidemic characteristics are predicted according to different rates of containment. The basic reproduction number is estimated to be 2.9949, with *CI*(2.6729–3.1485). Furthermore, a threshold value of containment rate, below which the epidemic duration is postponed, is determined.

**Conclusion:** Our findings show that the basic reproduction number reflects a high speed of spread of the epidemic. Furthermore, the compulsory containment can be efficient if more than 73% of population are confined. However, even with 90% of containment, the end-time is estimated to happen on July 4th which can be harmful and lead to consequent social-economic damages. Thus, containment need to be accompanied by other measures such as mass testing to reduce the size of asymptomatic population. Indeed, our sensitivity analysis investigation shows that the COVID-19 dynamics depends strongly on the asymptomatic duration as well as the contact and containment rates. Our results can help the Moroccan government to anticipate the spread of COVID-19 and avoid human loses and consequent social-economic damages as well.

## 1. Introduction

COVID-19 is an infectious disease caused by a novel betacoronavirus that primarily targets the human respiratory system **[12]**. Severity of COVID-19 symptoms can range from very mild to severe. Indeed, some people generally have mild to moderate respiratory illness and recover without requiring special treatment **[32]**. Others, like older adults and people with existing chronic medical conditions, are more likely to develop serious illness and have a high risk of death **[32]**. Human-to-human transmission of disease occurs via direct contact with the droplets saliva, discharge from the nose of an infectious person's coughs, sneezes or through contaminated objects and surfaces [32, 12]. The novel coronavirus was first emerged in the Chinese city of Wuhan in December 2019. In a few months, the virus rapidly spread throughout China and was exported to 114 countries around the world **[12, 35]**. The worldwide number of humans diagnosed with COVID-19 has surpassed 118,000 and nearly 4,300 people have died **[1]**. On March 11, 2020, the World Health Organization (WHO) has officially declared the novel coronavirus outbreak a pandemic **[2]**. To the best of our knowledge, no specific vaccine or antiviral exist for COVID-19 up to date, but there are many ongoing clinical trials evaluating potential treatments [14]. Meanwhile, classical public health measures such as isolation and quarantine, social distancing and community containment were adopted by many countries to prevent dissemination of the disease within their populations and to curb the epidemic [1, 35]. The containment measure, defined as an intervention applied to an entire community in order to lower intermixing of unreported infectious individuals with susceptibles as well as the spread of the virus [35], ranges from social distancing to community-use of face masks, including locking entire cities or areas.

Morocco is one of the countries affected by COVID-19. As of April 09,2020, the total number of confirmed cases in Morocco was reported to be 1374 infectious and 97 deaths [26]. The first case of Covid-19 infection was reported on March 02, 2020 in Casablanca city. The virus was imported from Italy by a Moroccan individual [16]. A few days later, many travelers from Italy, France or Spain have tested positive for Covid-19 in different cities of the country such as Casablanca, Marrakesh and Fez. Patients received care through isolation in hospitals and all people who have been in contact with them or, otherwise, high-risk for becoming ill are identified and tested [17, 18, 19, 20, 21, 22]. Since seven imported cases have emerged, Moroccan government decided to suspend flights to neighboring countries affected by COVID-19 such as Italy, Spain, Algeria and France [29]. However, reported cases continued to grow with eight new imported cases and a first confirmed local case appeared on March 14 [23]. The epidemic spreads to other Moroccan cities such as Tetouan, Rabat and Khouribga and, therefore, Morocco rapidly responded to the COVID-19 infection and the Moroccan authorities implemented a series of control measures to limit person-to-person transmission. The Ministry of interior has decided to close all schools of the country and promotes online interactive learning as an alternative [28]. Movements of Moroccan people are restricted to their living space and all gathering of more than fifty persons has become prohibited [29]. Mosques and non-essential common areas are closed except for pharmacies and stores selling necessary goods to citizen. Moreover, all international flights are suspended and all public events are canceled [29]. On March 19, the government decreed a state of health emergency and the compulsory containment of population has been declared [27]. However, a part of the population continued to work to supply the basic necessities to the confined population. Every day, infectious individuals are reported by the Ministry of Health and all individuals that have a close contact with them are identified and quarantined. Unfortunately, the unreported and asymptomatic infectious cases may exist within confined or working people and constitute a source of contamination of susceptible individuals like family members, co-workers, sellers, etc. Despite being the most affected country in the world by the disease, China is the first country that has succeeded in mitigating the progression of COVID-19 by a stringent containment of the population [33, 36]. Others countries such as Morocco still continue their fight to curb the growth in the number of daily infectious cases. Up to date, the number of confirmed cases of COVID-19 continues to increase and changes on a daily basis. The efficacy of containment and how long it must be maintained is still unknown. The longer the period of containment, the more serious consequences on the socioeconomic situation of the country will be. In addition, the short peak period of the epidemic and large notified cases prevent health care teams from adequately preparing for and responding to the huge load of patients. Thus, lowering the peak size and the postponing the peak time of the reported cases are of major interest.

In this study, using the reported cumulative confirmed cases in Morocco from March 2nd, 2020 to April 9, 2020 (See Table 1), we propose mathematical model, adapted to the Moroccan situation, to evaluate the efficacy of compulsory containment imposed by the government on March 20 in order to limit the disease transmission through Moroccan territory. We used this model to estimate the the basic reproduction number and determine the impact of containment on the peak time, epidemic duration, peak size and final size of the epidemic. This work can help to enhance understanding of the evolving of infection and to adjust the measures of containment in Morocco.

## 2. Methods

### 2.1. Data collection

The data of reported symptomatic infectious cases is collected each day at 11 pm from the official Coronavirus Portal of Morocco[26]. Data information covers the cumulative number of reported cases from March 2nd to April 9th. The data from March 2nd to March 20 are used to estimate the basic reproduction number and adjust the model (2.1) to become closer to reality, while data from March 21 to April 9 are used to adjust model (2.2) and estimate the containment rate during this period of containment.

**Table 1:** Cumulative daily reported case data from March 2nd, 2020 to April 9, 2020.

|  |  |  |  |  |  |  |  |  |  |  |
| --- | --- | --- | --- | --- | --- | --- | --- | --- | --- | --- |
| Date | 02/03 | 03/03 | 04/03 | 05/03 | 06/03 | 07/03 | 08/03 | 09/03 | 10/03 | 11/03 |
|  | 1 | 1 | 2 | 2 | 2 | 2 | 2 | 2 | 3 | 6 |
| Date | 12/03 | 13/03 | 14/03 | 15/03 | 16/03 | 17/03 | 18/03 | 19/03 | 20/03 | 21/03 |
|  | 6 | 8 | 17 | 28 | 37 | 44 | 54 | 63 | 86 | 96 |
| Date | 22/03 | 23/03 | 24/03 | 25/03 | 26/03 | 27/03 | 28/03 | 29/03 | 30/03 | 31/03 |
|  | 115 | 143 | 170 | 225 | 275 | 345 | 402 | 479 | 556 | 617 |
| Date | 01/04 | 02/04 | 03/04 | 04/04 | 05/04 | 06/04 | 07/04 | 08/04 | 09/04 |  |
|  | 654 | 708 | 791 | 911 | 1021 | 1120 | 1184 | 1275 | 1374 |  |

### 2.2. Model development

#### The model without containment

The population considered in our basic model, as shown in Fig. 2.1, is stratified into four disease status. Individuals are classified as susceptible 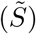, asymptomatic infectious (*Ã*), unreported symptomatic infectious (*Ĩ_u_*) and reported symptomatic infectious (*Ĩ_r_*).

**Figure 2.1:**
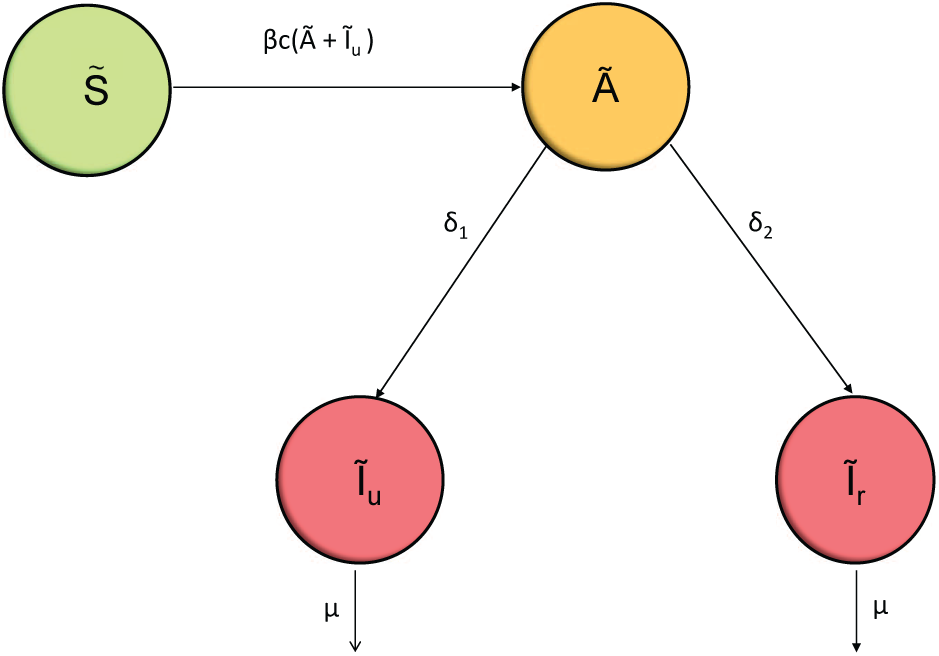
Basic 2019-nCoV transmission diagram

**Figure 2.2:**
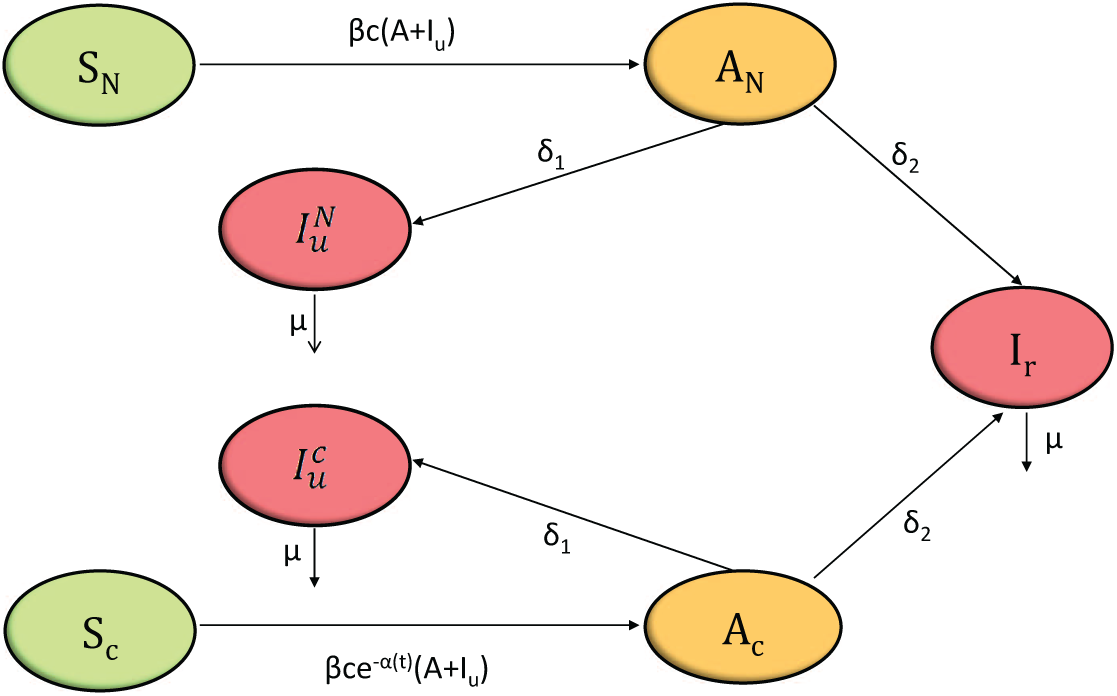
2019-nCoV transmission diagram with containment

Here, we assume that reported symptomatic infectious individuals are hospitalized and can not contact susceptibles anymore. We assume also, as confirmed by Rothe et al. [13], that asymptomatic individuals can infect susceptible individuals through an effective contact. Furthermore, MacIntyre in [15] proved that asymptomatic and symptomatic infectious individuals share the same infection probability. Taking account of the previous assumptions, the dynamics of COVID-19 can be described as follows: Susceptibles (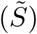) contacted with either unreported symptomatic (*Ĩ_u_*) or asymptomatic infectious individuals (*Ã*), at an effective contact rate, *c*, are infected with infection probability, *β*, and move to the asymptomatic infectious class (*Ã*). After an average period 1/*δ* days the asymptomatic infectious individuals (*Ã*) become symptomatic and proceed either to the unreported symptomatic infectious (*Ĩ_u_*), at rate *δ*_1_, or to the reported symptomatic infectious (*Ĩ_r_*) at rate *δ*_2_ with *δ* = *δ*_1_ + *δ*_2_. Once becoming symptomatic, individuals of class *Ĩ_u_* and *Ĩ_r_* remain asymptomatic for 1/*μ* days on average before they are recovered.

Since the containment measure started 19 days since the first reported case then the model equations without containment is defined for 0 ≤ *t <t*_0_:= 19 as follows

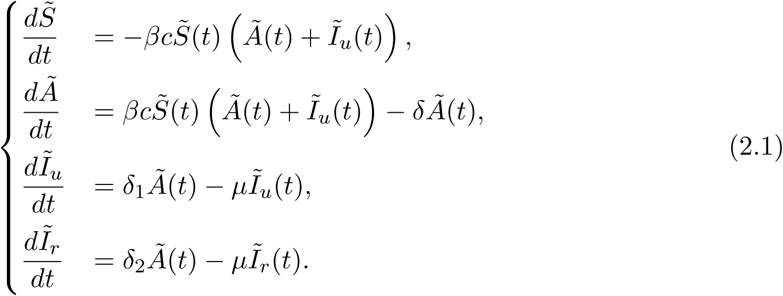

Initial data (states at *t* = 0 which correspond to March 2nd, 2020) of system (2.1) are estimated in Section 2.4.

#### The model with containment

Since the main purpose of this work is to study the spread of the COVID-19 across Morocco after the imposed measures made by the Moroccan government on March 21, 2020, the population considered in the basic model (2.1) will be further stratified according to unconfined susceptible individuals (*S_N_*), unconfined asymptomatic infectious individuals (*A_N_*), unconfined unreported symptomatic infectious individuals 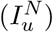, confined susceptible individuals (*S_c_*), confined asymptomatic infectious individuals (*A_c_*) and confined unreported symptomatic infectious individuals 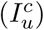. Here, we assume that confined asymptomatic and confined unreported individuals can still spread the virus to their families. Furthermore, since the Moroccan government starts to impose public major measures, and taking account the fact that Moroccan individuals gradually began to reduce their contact with the people nearby, we assume that the contact rate changes from a constant rate to an exponentially decreasing rate, *cl*(*t*), with time. It is meaningful to assume that the only subpopulation that will not be confined is the reported symptomatic infectious subpopulation.

The model with containment control will given, for *t* ≥ *t*_0_ days, by the following equations

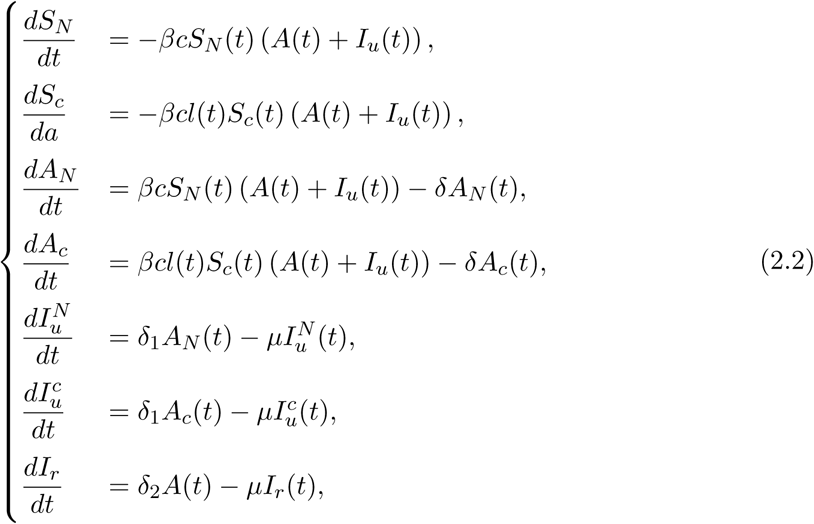

where *cl*(*t*) = *ce*^-^*^α^*^(^*^t^*^−19)^ is the time-dependent contact rate, *A*(*t*) = *A_N_*(*t*) + *A_c_*(*t*) is the total asymptomatic infectious population and 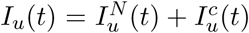 is the total unreported symptomatic infectious population. Moreover, if the containment rate of susceptible, asymptomatic infectious and unreported infectious subpopulations is denoted by *p*, then the new initial data for system (2.2) shall be given by 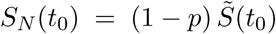, 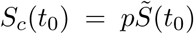, *A_N_*(*t*_0_) = (1 − *p*) *Ã*(*t*_0_), *A_c_*(*t*_0_) = *pÃ*(*t*_0_), 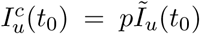, 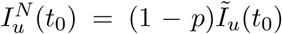 and *I_r_*(*t*_0_) = *Ĩ_r_*(*t*_0_).

### 2.3. The reproduction number

The basic reproduction number, *R*_0_, is the average number of secondary infections produced when one infectious individual is introduced into a host susceptible population. This quantity determines whether a given disease may spread, or die out in a population. To compute this number, we apply the next generation matrix method in [31]. Computation method of *R*_0_ is presented in the Appendix.

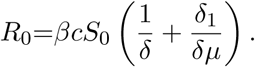

Here, *R*_0_ can be explained as follows: Assume that one asymptomatic infectious individual is introduced into the susceptible population. This asymptomatic individual produces, on average, 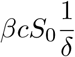 asymptomatic individuals during his average lifespan 1/*δ*. These asymptomatic individuals then become unreported symptomatic infectious individuals over their lifespan 1/*δ* at a rate *δ*_1_ and then each infectious symptomatic produces, on average, 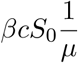asymptomatic individuals during his lifespan 1/*μ*.

The control reproduction number, *R_c_*, is an important value, used to determine whether a control policy, such as containment in our case, will be efficient to decrease the number of secondary infections to be less than one. Here, we compute the control reproduction number related to the early stage of the containment (During the first 20 days starting from the first day of containment). Computation method of *R_c_* is presented in the Appendix.

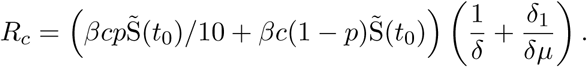

### 2.4. Parameter estimation

The parameter estimation is a crucial step of our study since the epidemiologically relevant choice of the parameters must establish and confirm the observed dynamics of the infection during the onset of the epidemic. Table 2 gives the values of the parameters used in the model. In our simulations, the the total population of the Moroccan Kingdom, 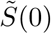, is chosen based on estimates from [5], the asymptomatic duration, 1/*δ*, based on estimates from [26] and the symptomatic duration, 1/*μ*, based on estimates from [34]. Note that the data of reported cases used to estimate the model parameters was carried out before containment and during the epidemic period. Thus, we assume that the contact rate, *c*, since the first reported case and before the first day of containment is around 10 contacts per person in average [9]. Furthermore, we assume that a likely confined individual can only go out once every 2 weeks on average to do the necessary shopping while an individual who cannot respect containment, due to a job of paramount importance, must go 5 days a week to work. Thus, it is meaningful to assume that, during the short time from March 21 to April 9, the average contact rate satisfy the relation 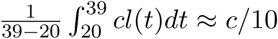. Thus, we assume that *α* ≈ 0.078. The other parameters and initial data are estimated as follows:

Since the first and the only infectious symptomatic individual is reported on March 2nd, 2020, which corresponds to *t* = 0, then 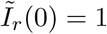. For the estimation of *β, Ã*(0) and *Ĩ*(0) we will use the data of cumulative reported cases collected from March 2nd to March 20 (before the start of containment) in Table (1) and we follow the procedure by [8]. The cumulative reported infectious population is given, for *t* ≥ 0, by 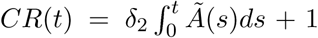. It is obvious that cumulative reported infectious population increases slowly and then accelerates rapidly with time. Hence, we will use exponential regression with 95% of confidence level to find an exponential function that best fits the data, from March 2nd to March 20th, in Table (1). We found that exponential model given by *be^at^* with *a* = 0.263 with confidence interval *CI* (0.229 – 0.297) and *b* = 0.507 with *CI* (0.3444 – 0.7475) fits well the data with a correlation coefficient given by *R* = 0.97. It follows from 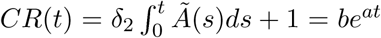
that

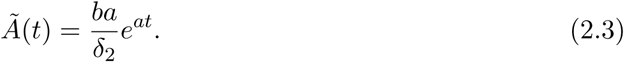

Since the initial susceptible population is not dramatically affected in the early phase of the epidemic, we will assume that 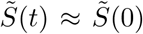. Let 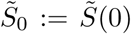, *Ã*(0):= *Ã*_0_ and *Ĩ_u_*(*0*):= *Ĩ*_0_. From the second equation of system (2.1) and using (2.3) we obtain

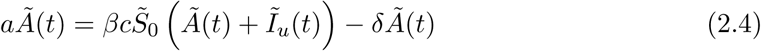

and

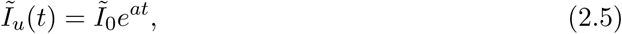

where

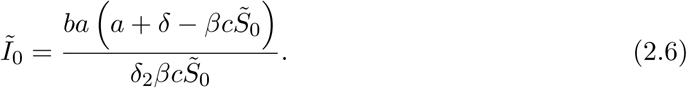

Now, using equations (2.4) and (2.5) and the third equation of system (2.1), we obtain after simplification

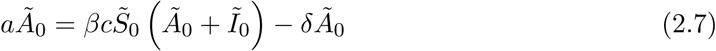

and

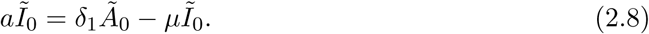

Solving equations (2.6), (2.7) and (2.8) for *β* and *Ã*_0_ lead to

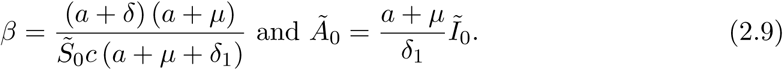

**Table 2:**
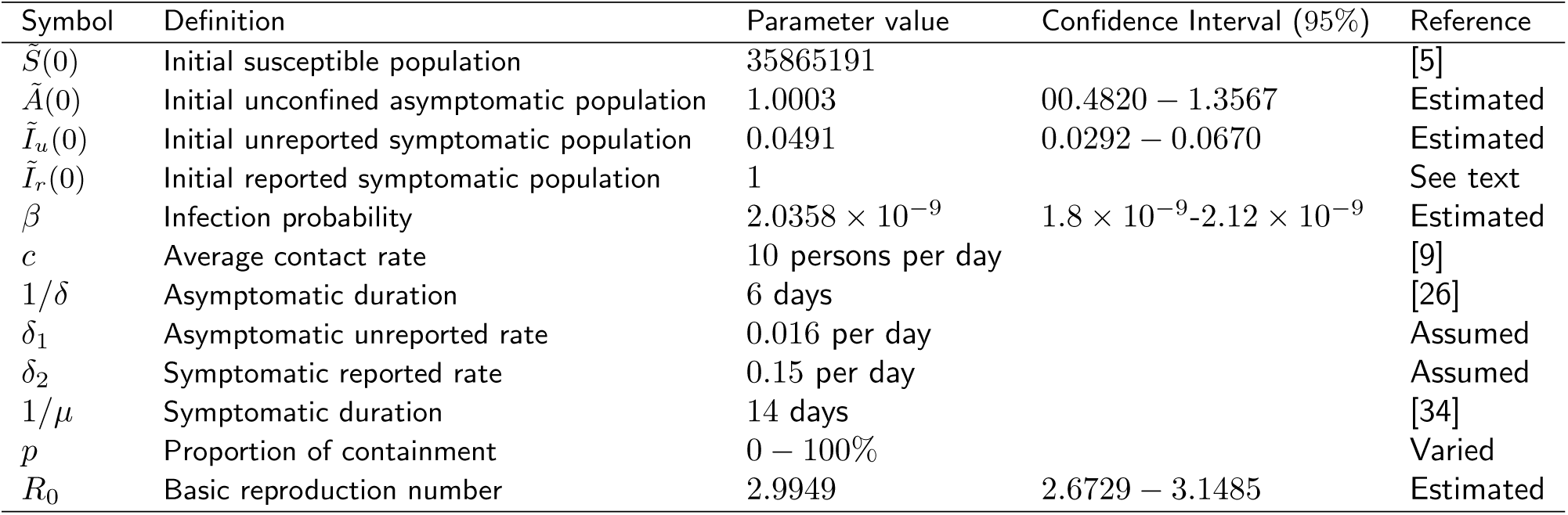
Parameter definitions and values of COVID-19 model.

## 3. Results and discussion

The government have deployed a series of severe control measures to limit transmission of COVID-19 across Morocco. The Moroccan people entered compulsory containment 19 days since the first infectious case has been confirmed. In this study we adopted a deterministic mathematical model of COVID-19 dynamics which took into account the Moroccan containment strategy used to control and eradicate the disease in the country. We used reported infectious case data, from March 2nd to April 9th, 2020, provided by the Health Ministry of Morocco to parameterize the model. On the one hand, as shown in Fig. 3.1, our simulations shows that our model fit well the cumulative data of reported infectious cases giving in Table 1, under the absence of compulsory containment conditions.

**Figure 3.1:**
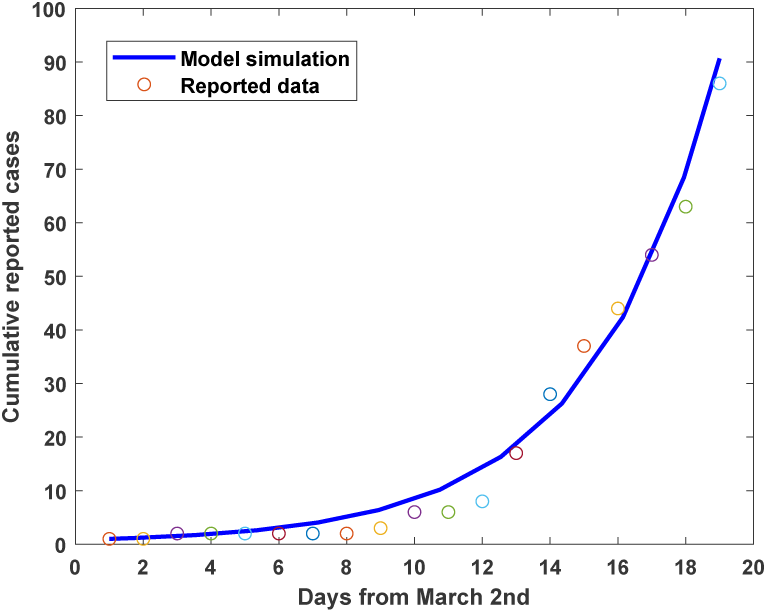
Comparison between time series of cumulative reported cases for model (2.1), with *a* = 0.296 and *b* = 0.555, and reported data of cumulative confirmed cases of COVID-19 from March 2nd to March 20, 2020. The dots are the day by day reported data in Table 1. The blue solid line is the fitting curve of model (2.1).

We estimated the basic reproductive number *R*_0_ at 2.9949 with *CI* (2.6729 – 3.1485) reflecting a high rate of COVID-19 transmission. This result is in agreement with the basic reproduction number estimated and reported by Liu et al. [7] using twelve values of *R*_0_ determined by many authors as a sample. According to the variability of the mathematical models used, Liu et al. estimated that *R*_0_ is expected to be between 2.79 and 3.28, which exceeds the basic reproduction number reported by WHO [30] ranging from 1.4 to 2.5.

On the other hand, to be as close as possible to reality, we used the cumulative reported case data from March 21th to April 09th and we applied the same method in Section 2.4 to estimate the containment rate *p* during these first 20 days of containment (See appendix 4). The Fig. 3.2 showed that our model is able to predict the provided real reported data during the first 20 days past after the beginning of containment.

**Figure 3.2:**
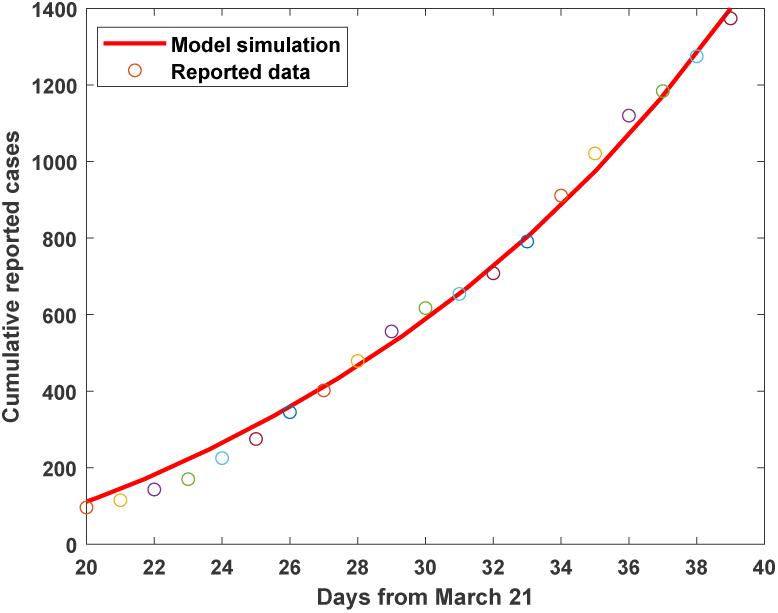
Comparison between time series of cumulative reported cases for model (2.2) with *p =* 0.55 and reported data of cumulative number of confirmed cases of COVID-19 from March 21 to April 9, 2020. The dots are the day by day reported data in Table 1. The red solid line is the fitting curve of model (2.2).

We estimated that only 55% of Moroccan population including susceptible, asymptomatic and unreported infectious individuals are confined during this period. This weak proportion can be explained by an adaptation time taken by citizens to the new living conditions at the beginning of containment. Unlike the Chinese people which has learned lessons from past epidemics, it is the first time that the Moroccan people face an epidemic and it is difficult to change their behavior spontaneously. Fortunately, facing the gravity of the situation, the Moroccan people subsequently showed themselves to be able quickly to adapt to the compulsory containment measures. The rate of containment certainly increased over time and Morocco can halt the epidemic with far fewer victims. However, according to our simulations, if Morocco continued with this containment rate (*p* = 0.55), the cumulative number of reported infectious cases would reach 93,906,000 reported cases. Furthermore, the peak-size of the number of reported infectious cases would be 120700 with a peak-time of 183 days. The last infectious case will be reported after more than one year (Fig. 3.3).

**Figure 3.3:**
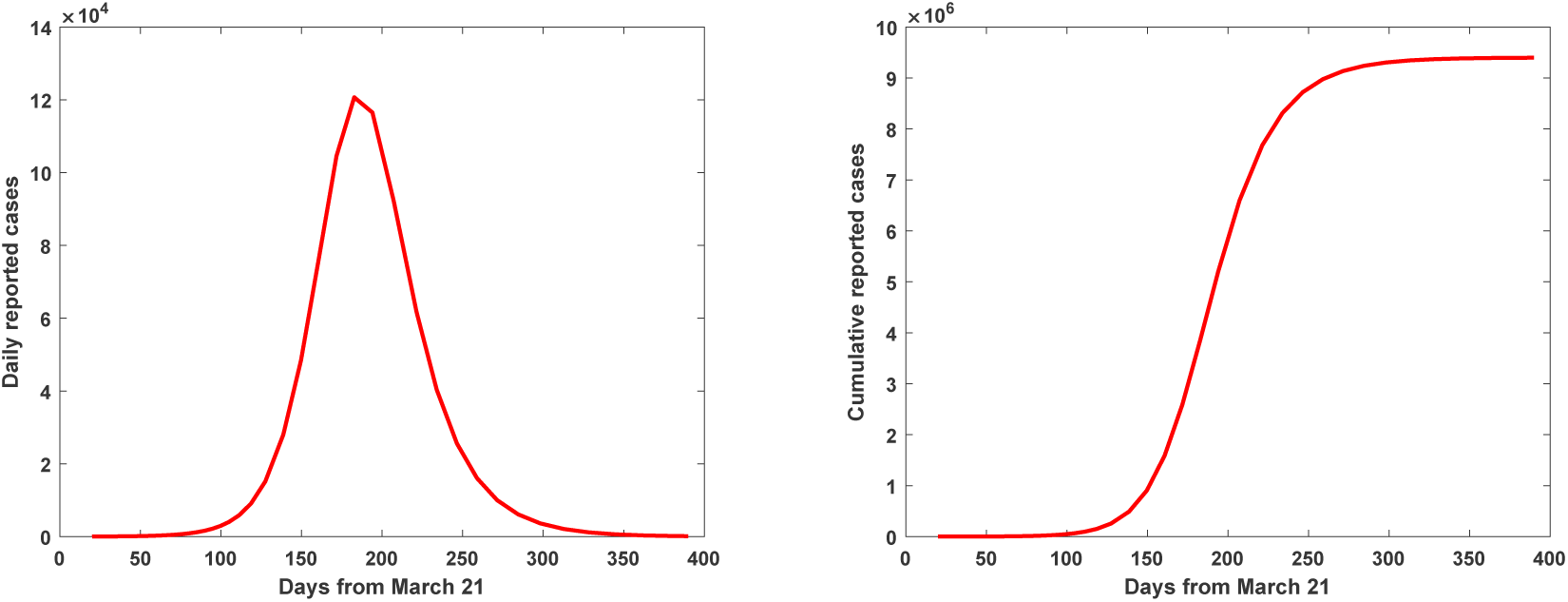
Time series plot for model (2.2), starting from March 21, 2020, with *p* = 0.55. Left panel corresponds the numbers of confirmed symptomatic infectious individuals. Right panel corresponds the numbers of cumulative confirmed symptomatic infectious individuals.

These results are horrifying and no country in the world could control a major wave of infectious cases. For practical reasons, it is imperative to reduce the peak-value so as not to exceed the limit capacity of hospital beds and lead the country towards an uncontrollable dramatic situation. It is also better to delay the peak appearance time to allow official health structures to prepare for such a high level of emergency. Increasing the containment rate *p* from 0.73 to 0.90 lead to a rapid saturation of cumulative number of reported infectious cases, ranging from 16950 to 7558 (Fig. 3.4). Furthermore, as shown in Fig. 3.4, the peak times of reported infectious cases decreased slightly while the epidemic duration decreased significantly from 180 to 125 days and the peak-sizes decreased from 338 to 188 cases (See Table 3). If Moroccan government succeeded to contain more than 90% of population, then the peak size of infectious cases would be less than 188 reported cases on April, 27th, 2020.

**Figure 3.4:**
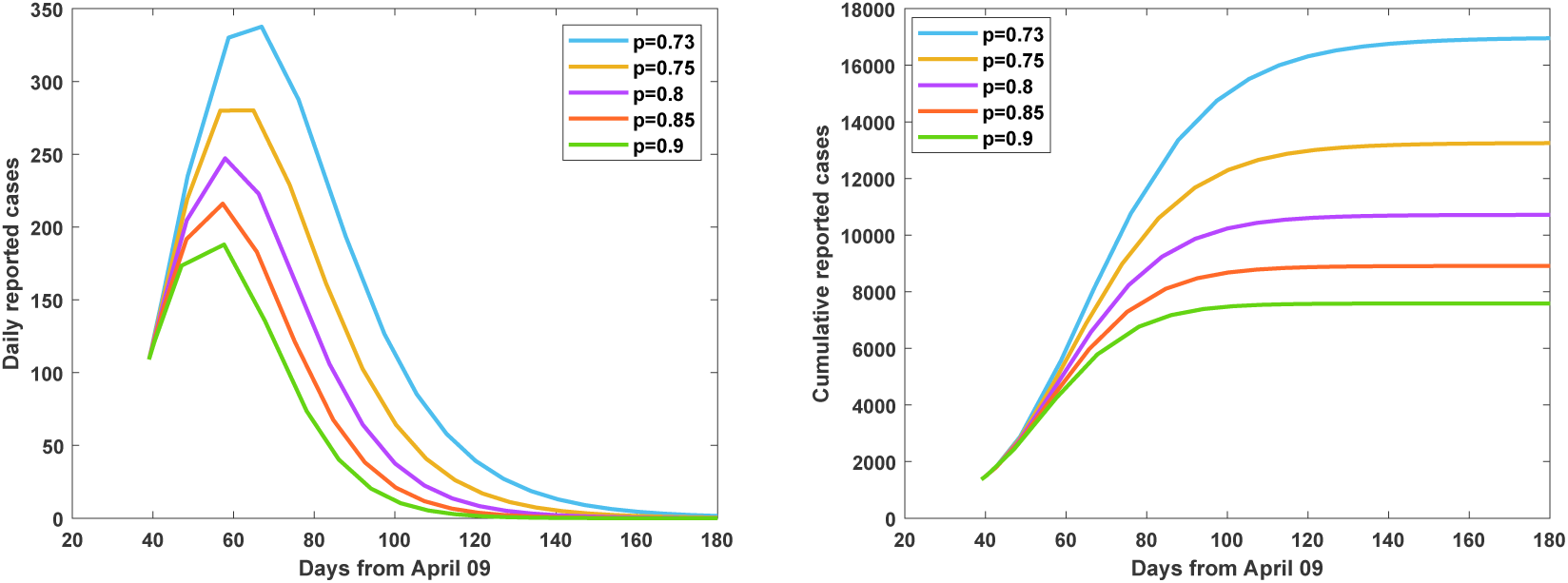
Time series plot for model (2.2), starting from April 9,2020, of the numbers of reported individuals change using different containment rates (*p* ∈ {0.73, 0.75, 0.8, 0.85, 0.9}). Left panel corresponds the numbers of daily reported individuals. Right panel corresponds the numbers of daily cumulative reported individuals.

However, these results could be overestimated or underestimated since many epidemiological factors are still unknown and are subject of study by researchers. Up to date, Morocco still did not reach the epidemic peak yet. Furthermore, if the containment period will last so long, Morocco must combine its actual strategy with the mass-testing or the contact-tracing strategies which allow finding and controlling the suspected cases through the susceptible population [10, 6]. For example, to the best of our knowledge, South Korea is the only country that has succeeded to greatly slow its epidemic without resorting to drastic containment [3].

**Table 3:** The effect of containment rate on the peak size, peak time, epidemic duration since March 2*nd*, 2020 and final cumulative cases

| Containment rate | Peak size | Peak time | Epidemic duration | Final cumulative cases |
| --- | --- | --- | --- | --- |
| $p = 0.73$ | 338 | May 7 | 180 | 16950 |
| $p = 0.75$ | 280 | From April 27 to May 5th | 167 | 13240 |
| $p = 0.8$ | 247 | April 28 | 151 | 10710 |
| $p = 0.85$ | 216 | April 27 | 136 | 8908 |
| $p = 0.9$ | 188 | April 27 | 125 | 7558 |

The increase of the containment rate does not significantly affect the peak-time but acted differently on epidemic duration (see Fig. 3.4). Indeed, we found that, by increasing the containment rate, there exists a threshold control reproduction number *R_c_* = 1 corresponding to a threshold containment rate *p*\* = 0.73, below which the epidemic end-time is postponed (Fig. 3.5) and above which the epidemic end-time is advanced (Fig. 3.4). This phenomenon can be explained as follows: In the absence of containment, the whole population becomes infected within a short time. However, by increasing the rate of containment gradually to the threshold *p*\*, the daily number of infectious individuals becomes lower and continues to significantly infect the susceptible individuals, which increases the duration of the epidemic. Moreover, if the containment rate passe the threshold, the containment becomes efficient since the daily number of infectious individuals significantly diminish, which in turn decrease the number of new infected and, consequently, shorten the duration of the epidemic.

**Figure 3.5:**
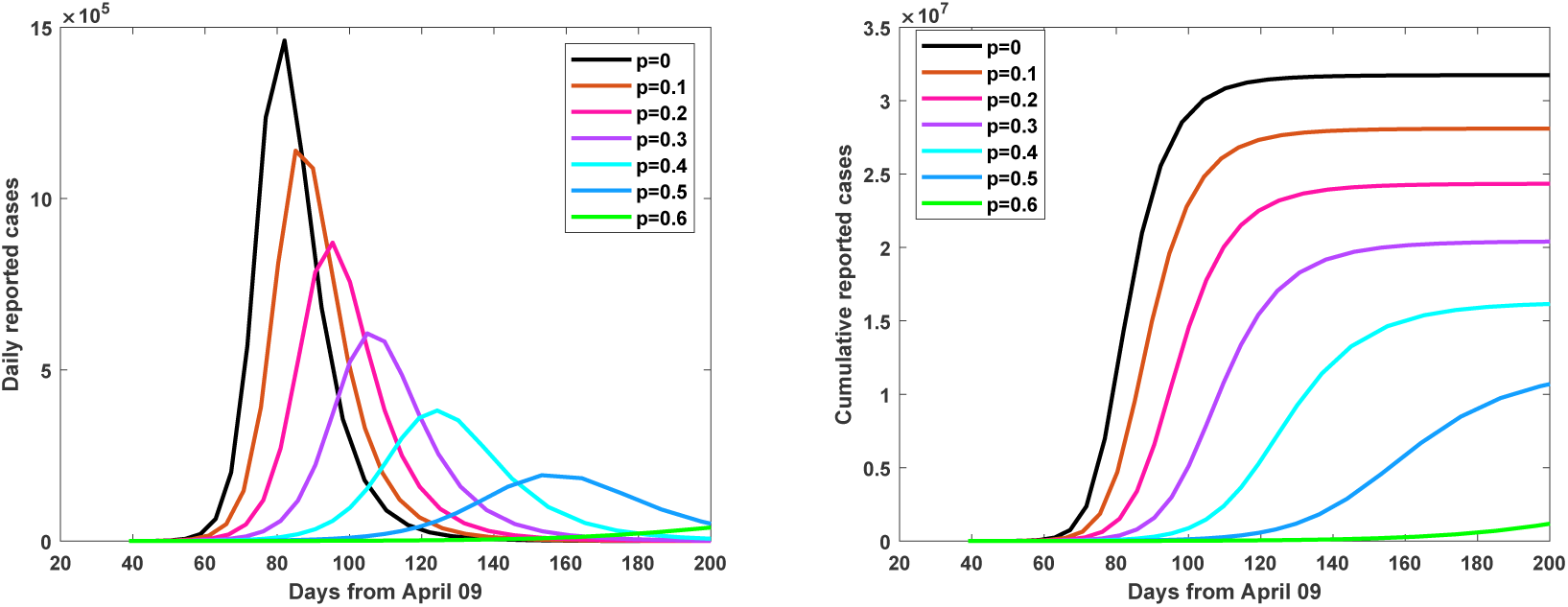
Time series plot for model (2.2), starting from April 9, 2020, of the numbers of reported individuals change using different containment rates (*p* ∈ {0.1, 0.2, 0.3, 0.4, 0.5, 0.6}). Left panel corresponds the numbers of daily reported individuals. Right panel corresponds the numbers of daily cumulative reported individuals.

Finally, the study of sensitivity analysis supported the results above and found that the containment rate has significant impact with correlation coefficient PRCC=−0.8. In general, the only three parameters playing a major role on the model dynamic are the contact rate, the containment rate and the asymptomatic duration (Fig. 3.6).

**Figure 3.6:**
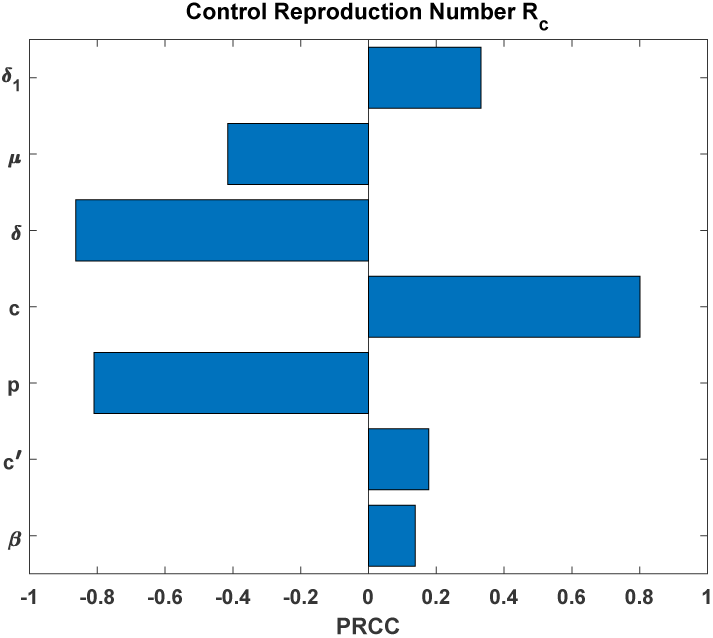
Sensitivity analysis. The partial rank correlation coefficients are shown using the control reproduction number.

However, our sensitivity analysis study show that the asymptomatic duration have a greater impact, with correlation coefficient PRCC=0.863, and consequently, can play a major role in COVID-19 spread across Morocco. Indeed, it has been shown that younger age individuals are the most contributor of silent transmission of COVID-19 to older family members [15], which could be the case in Morocco since 26% of the Moroccan community are less then 15 years old [4]. Fortunately, the Moroccan government have decided to close schools earlier to avoid such a critical epidemic consequences.

## 4. Conclusions

The COVID-19 infection was primary imported to Morocco from European countries. The Moroccan government have deployed a series of rigorous control measures to limit person-to-person transmission of COVID-19 and imposed the compulsory containment of population on March 21, 2020. Our modelling results of these strategies proved that containment played a major role in the spread limitation of infection and seems to be influenced by the rate of asymptomatic individuals. A containment of 90% of Moroccan population including susceptible, unreported symptomatic and asymptomatic infectious individuals can lead to the elimination of COVID-19 probably around July 4th, 2020. This deadline is only approximate, without commitment, due the lack of accurate health data in Morocco such as the symptomatic period, the duration from onset symptoms to recovery or death etc. As long as more data becomes available, our model can be comfortably updated. Furthermore, our model does not consider the influence of the age of population which seems to be very important since younger age individuals contributor greatly of silent transmission of COVID-19. Furthermore, antiviral treatment or vaccine and the climatic factors on the dynamic of the disease should be included in the model. An extension of the current model may be to assess the impact of the contact-tracing and mass testing strategies on COVID-19 in Morocco. Actually, the Moroccan government has established several diagnostic centers in several cities to increase the number of COVID-19 tests and decrease the harmful effect of asymptomatic infectious cases.

## Data Availability

All data referred to in the manuscript are available.

https://www.covidmaroc.ma

## Appendix

### Basic reproduction number

The linearized equation related to infectious individuals of system (2.1) is given by

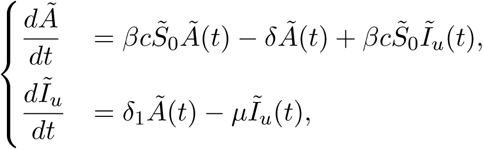

and the associated Jacobian matrix is given by

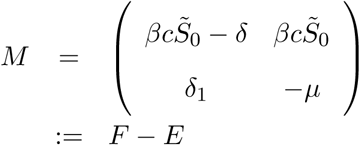

where

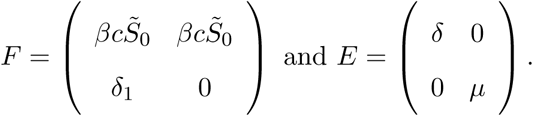

Therefore, 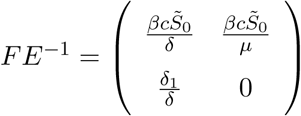 and *R*_0_ is its spectral radius which is given

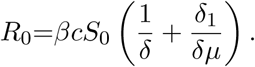

### Control reproduction number

Following the remark on the contact behaviour and the estimation given in Section 2.4 we will assume that, in average, *cl*(*t*) = *c*/10. The linearized system of the related to the disease is given by

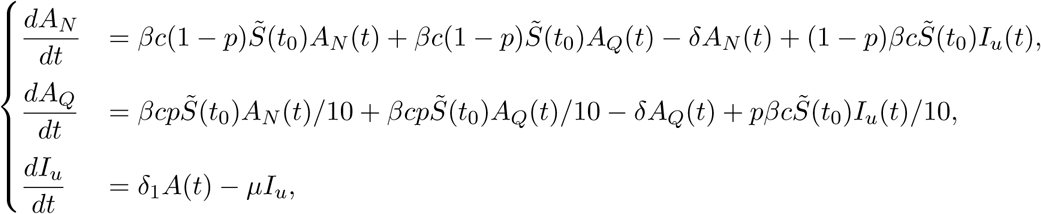

and the associated Jacobian matrix is given by

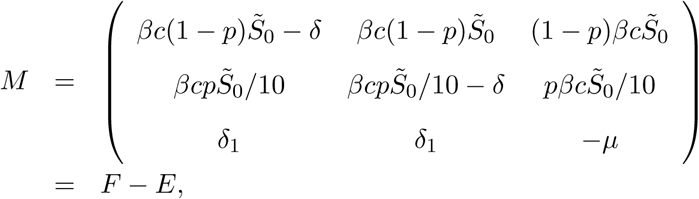

where

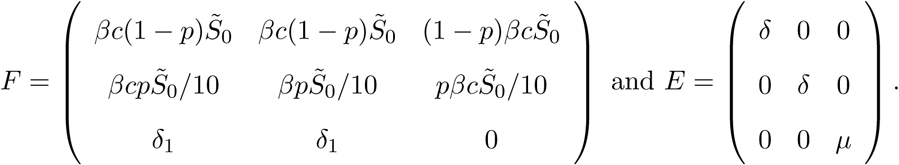

It follows that

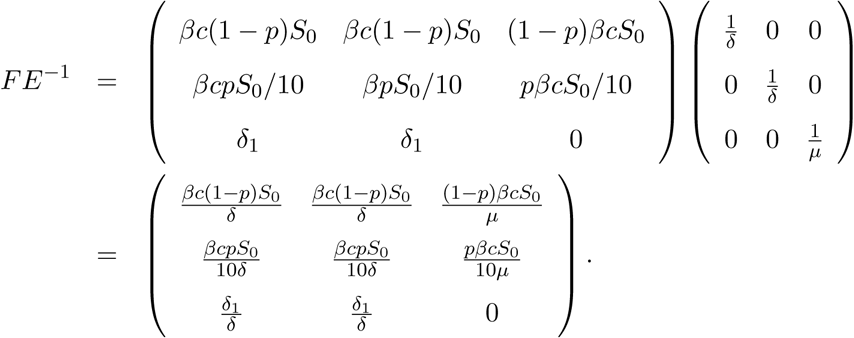

Consequently, the control reproduction number is given by

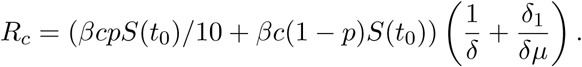

### Estimation of p between March 21 and April 9

The cumulative reported cases for *t* ≥ *t*_0_ is given by 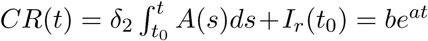. Here we will use once again the exponential regression with 95% of confidence level to find an exponential function that best fits the data, from March 21 to April 9, in Table (1). We found that exponential model given by *be^at^* with *a* = 0.139 with confidence interval *CI* (0.125 − 0.153) and *b* = 7.8303 with *CI* (5.176 − 11.858) fits well the data with a correlation coefficient given by *R* = 0.981. Differentiating the both terms of *CR*(*t*) leads to

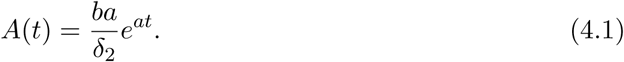

Since the initial susceptible population is not dramatically affected in the early phase of the epidemic, we will assume that 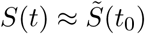. Adding the third and fourth equation of system (2.2) and using (4.1), we obtain

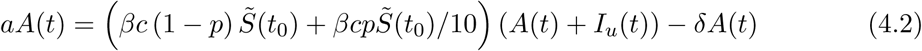

and

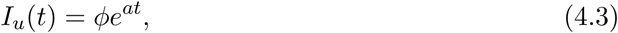

where

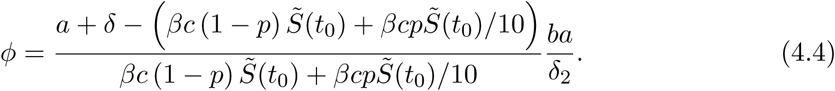

Now, using equations (4.2), (4.3) together with the the fifth and sixth equations of system (2.1), we obtain, for any *t* ≥ *t*_0_ such that *A*(*t*) ≠ 0,

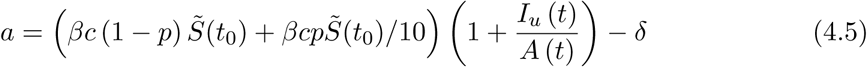

and

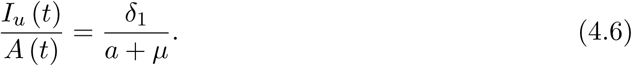

Solving equations (4.5) and (4.6) for *p* lead to

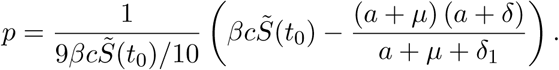

## Acknowledgments

The authors thank the anonymous referees, whose careful reading, insights, valuable comments, and suggestions significantly enabled us to improve the quality of the paper.

